# Leveraging genomic large language models to enhance causal genotype-brain-clinical pathways in Alzheimer’s disease

**DOI:** 10.1101/2024.10.03.24314824

**Authors:** Qiao Liu, Wanwen Zeng, Hongtu Zhu, Lexin Li, Wing Hung Wong, Alzheimer’s Disease Neuroimaging Initiative

**Affiliations:** Department of Statistics, Stanford University, Stanford, CA 94305, USA; Department of Biomedical Data Science, Stanford University, Stanford, CA 94305, USA; Department of Biostatistics, University of North Carolina at Chapel Hill, Chapel Hill, NC, USA; Biomedical Research Imaging Center, University of North Carolina, Chapel Hill, NC 27599, USA; Division of Biostatistics, University of California, Berkeley, CA 94720, USA; Bio-X Program, Stanford University, Stanford, CA 94305, USA

## Abstract

Genome-wide association studies (GWAS) have identified numerous Alzheimer’s disease (AD)- associated variants. However, how these variants contribute to the etiology of AD remains largely elusive. Recent advances in genomic large language models (LLMs) offer new opportunities to interpret the genetic variation observed in personal genome. In this study, we propose epiBrainLLM, a novel computational framework that leverages genomic LLM to enhance our understanding of the causal pathways from genotypes to brain measures to AD-related clinical phenotypes. epiBrainLLM will first convert the personal DNA sequence into a diverse set of genomic and epigenomic features using a pretrained genomic LLM and then use these features to further predict phenotypes. Across various experimental settings, epiBrainLLM significantly improves causal analysis compared to traditional genotype association approach. We conclude that epiBrainLLM provides a novel perspective for understanding the regulatory mechanisms underlying the AD disease etiology, potentially offering insights into complex disease mechanisms beyond AD.

## Introduction

Alzheimer’s disease (AD) has emerged as a global healthcare crisis^1^. According to a recent WHO report^2^, around 55 million people globally have dementia, with approximately 40 million of these cases (72.7%) being AD. Over 60% of those affected reside in low- and middle-income countries. The total number of people with AD is projected to rise to about 78 million by 2030 and potentially 139 million by 2050. Despite substantial investments, AD remains one of the most complex brain diseases with no cure or effective prevention strategy. Both the scientific community and healthcare sectors recognize the urgency to improve the understanding of the etiology of AD.

AD has a strong genetic basis with the heritability estimated to range from 60% to 80%^3,4^. A considerable number of AD-associated loci or genes have been identified by GWAS. For example, a recent large-scale GWAS study involving 111,326 AD cases and 677,663 controls discovered 75 risk loci, of which 42 were new at the time of analysis^5^. Despite the discovery of many genetic regions associated with AD risk, the identification of causal variants within these regions remains challenging^6,7^. Several recent studies predicted that there are about 100 to 10,000 causal variants contributing to AD while only a small fraction of them have been identified so far^8,9^. Importantly, GWAS studies on AD typically ignored the importance of regulatory mechanisms in gene expression and disease development^10^. Genetic variants can modulate disease risk through both direct effects on protein functions and indirect effects on regulatory activities, often involving the participation of *cis*-regulatory elements, such as promoters and enhancers, and *trans*-acting factors, such as transcription factors^11–15^. These *cis*- and *trans*-regulatory activities contribute significantly to the gene expression control and the genetic architecture of complex traits^16–18^. Failure to consider these regulatory mechanisms may result in an incomplete comprehension of AD etiology.

Decoding the functions of AD risk variants in the human genome is mainly hindered by the vast non-coding regions^19,20^. Consequently, AD research will benefit from novel computational methods that can infer how non-coding variants disrupt the underlying regulatory syntax of DNA, ultimately leading to dysfunctions of gene regulation that elevate disease risk. The inherent similarities between natural language and DNA sequences offer useful perspectives for understanding the information encoded in a personal genome^21^. Recently, transformer-based large language models (LLMs) have led to breakthroughs in various natural language processing (NLP) and computer vision tasks^22,23^, while presenting unprecedented opportunities to uncover the complex regulatory syntax of DNA, such as dissecting of the universal gene regulation rules and providing insights into understanding disease etiology. Genomic LLMs have already achieved state-of-the-art performance in a variety of regulatory genomic prediction tasks^24–27^. Integrating the genomic LLMs into AD research therefore has the potential to significantly advance our understanding of AD etiology.

In this study, we systematically leverage genomic LLM models to enhance our understanding of causal pathways from genotypes to brain imaging measures to AD-related clinical phenotypes. We consider various brain imaging measures derived from anatomical imaging data (e.g., MRI) and clinical phenotypes from clinical diagnoses (e.g., cognitive scores, and AD diagnosis status). Existing imaging genetics studies aim to understand the biological pathways through which genes affect brain imaging measures^28,29^. Moreover, most existing AD GWAS studies focus on identifying genetic variants that are statistically associated with AD disease. Here, we propose to investigate epigenomic features predicted by genomic LLMs underlying both brain imaging measures and AD-related clinical data. These features represent intermediate causal variables relevant to gene regulation, potentially revealing new insights into the mechanisms driving AD. Our new epiBrainLLM approach offers several advantages. First, genomic LLM converts personal DNA sequence into a set of personal genomic and epigenomic signals, including chromatin accessibility, transcription factor binding, and histone modifications, which considers the combinatorial effects of multiple variants, especially the rare or de novo variants, and their interactions within a large genomic region, providing a more holistic view of genetic influence on phenotypic traits. Second, our epiBrainLLM approach investigates genomic and epigenomic signals as intermediate features underlying various AD-related biomarkers (e.g., brain imaging measures) that reflects the pathological processes underlying the progression of AD. Third, our approach is able to amplify the genetic signals through transferring the knowledge learned from the pretrained genomic LLMs and integrating information from multiple genetic variants that affect the regulatory status of a genomic region.

We demonstrate the effectiveness of our approach through a series of experiments. First, we showed that our proposed epiBrainLLM can better associate each AD causal or risk gene with specific brain structure and function through different imaging phenotypes. Second, we illustrated that epiBrainLLM can better evaluate the contribution of each gene to a diverse set of AD clinical phenotypes. The top-ranked genes identified through our approach are highly consistent with the literature. The constructed maps could deepen our understanding of AD genetics-brain-clinical pathways information, providing a comprehensive picture of how genetic variants are causally related to brain imaging measures and AD clinical variables mediated by personal genomic and epigenomic features. We conclude that our new epiBrainLLM approach could shed light on understanding the pathophysiological processes in AD and facilitate the identification of new biological features, new prognostic/diagnostic markers and new therapeutic targets for AD through translational genomics.

## Results

### Overview of our study design

We propose epiBrainLLM, a novel computational framework that leverages genomic LLM to enhance our understanding of the causal pathways from genotypes to brain measures to AD-related clinical phenotypes, providing new insights beyond the traditional GWAS and imaging genetics studies. Figure 1A illustrates the pipeline of our proposed epiBrainLLM approach. Unlike conventional methods that directly associate genetic features with phenotypes, our new approach epiBrainLLM, empowered by the genomic LLM, amplifies the genetic signal by integrating information from numerous genetic variants affecting the regulatory status of a genomic region. The pretrained genomic LLM first transforms the personal genome DNA sequence from the above genomic region into personal genomic LLM features, which include genomic and epigenomic signals across different cellular contexts. These genomic LLM predicted features serve as intermediate causal variables relevant to gene regulation, potentially revealing new insights into the mechanisms driving AD. We then perform a regression analysis (or classification) between these genomic LLM imputed features and phenotype of interest (either imaging phenotypes or AD clinical phenotypes). Since the predicted intermediate features are not correlated with the confounders (factors that affect both the actual intermediate features and the phenotype), the relationship between the intermediate features and the phenotype variable in the second regression step can be interpreted as causal or less biased^30,31^. The signal amplification powered by genomic LLM alleviates the small sample size issue and provide a stronger detection power in discovering genotype-brain-clinical pathways in AD. One notable characteristic of our proposed approach lies in its ability to handle the whole genome sequencing (WGS) data, which is becoming increasingly available^32^. The WGS data enables our approach to consider the combinatorial effects of multiple genetic variants, including rare variants with low frequency that may have substantial impacts on disease risk^33,34^. More details for the WGS data preprocessing pipeline (Figure 1B) and magnetic resonance imaging (MRI) data preprocessing pipeline (Figure 1C) are given in the Methods section. Our approach has the potential to provide a comprehensive understanding of how genetic variants influence AD imaging traits and disease risk through genotype-brain-clinical pathways.

**Figure. 1.**
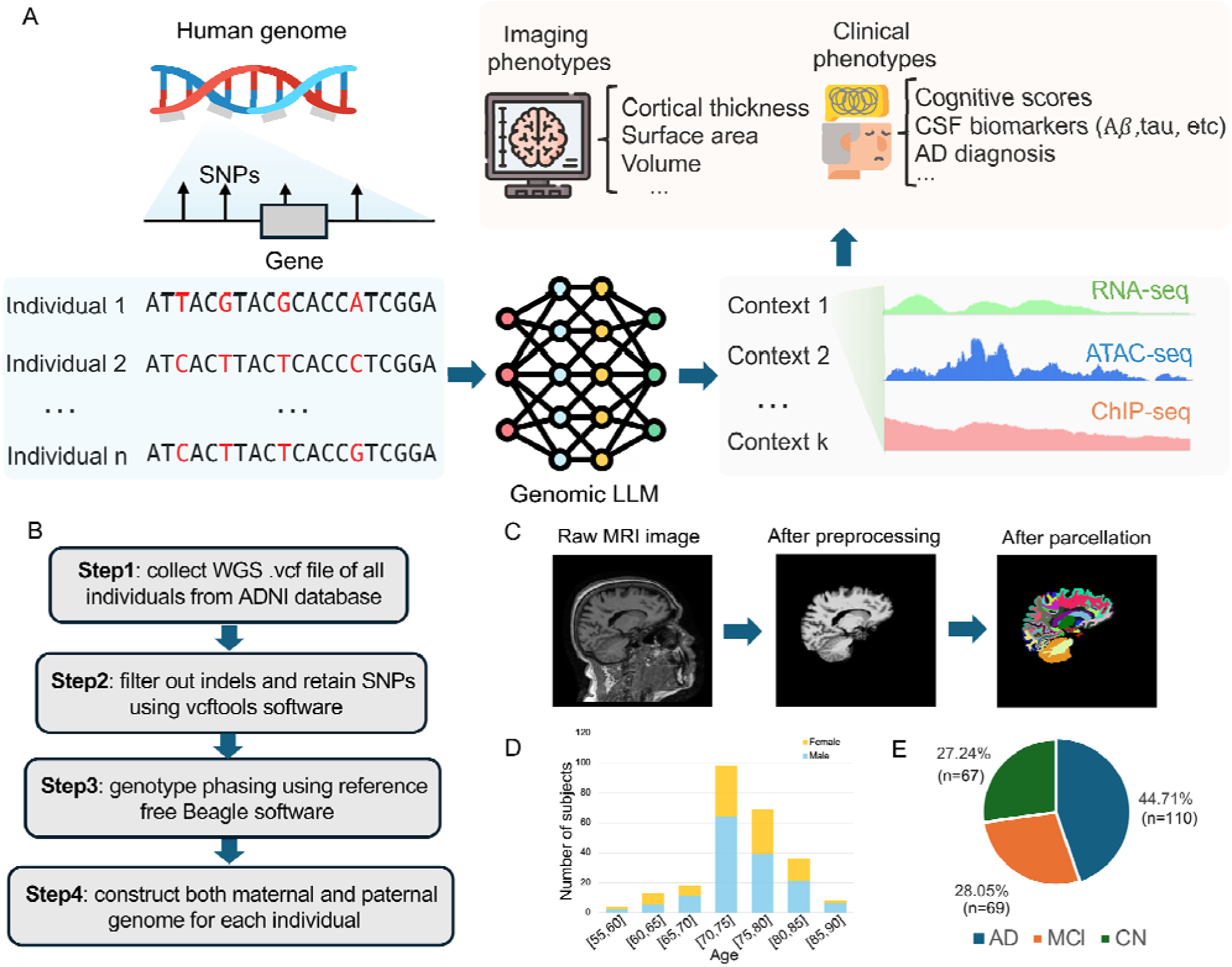
The overview of our study design. (A) Our approach pipeline epiBrainLLM first transforms personal genome sequences (e.g., whole genome sequencing) into genomic L M features and then associate these features to different phenotypes. (B) The pipeline for preprocessing the whole genome sequencing (WGS) data for use in Step A. (C) The pipeline for preprocessing the MRI imaging data of human brain. (D) The age and gender distribution of the participants with both WGS and longitudinal image data. (E) The clinical diagnosis AD status of the participants with both WGS and longitudinal image data.

To demonstrate the effectiveness of our proposed approach epiBrainLLM, we downloaded the WGS data from 808 individuals and the structure MRI data across three time points (baseline, 6^th^ month, and 12^th^ month) of 639 individuals from Alzheimer’s disease neuroimaging initiative (ANDI) database^35^. A subset of 246 individuals with both WGS and imaging data were obtained, which were used for all the imaging and longitudinal analysis. The age/gender distribution for these individuals is given in Figure 1D and the corresponding clinical AD diagnosis is given in Figure 1E. In the clinical phenotype analysis, including the protein biomarkers in cerebrospinal fluid (CSF), there are 785 individuals with both WGS and clinical phenotypic data, including protein biomarkers and cognitive scores.

### Investigating the causal relationship between gene and brain ROI

The structural changes in the human brain are known to be both phenotypically and genetically associated with neurodegenerative diseases, such as Alzheimer’s disease^36^. Individual human brain structural features, such as cortical thickness, surface area, and volume can be measured quantitatively by structural magnetic resonance imaging (sMRI). These whole brain MRI scans are processed and annotated to identify pre-defined regions of interest (ROIs). Using our association approach empowered by the genomic LLM (Figure 1A), we are able to test the association of a gene-centric genomic region consisting of multiple genetic variants to each of the ROI in the human brain. Since the sample size is not large enough to do a genome-wide scan of associations, we focused on a set of genes known to be associated to AD, and used them to compare the strength of association signal offered by our epiBrainLLM and the traditional genotype-based approach.

We first collected 16 AD causal genes and additional nearest genes from 56 AD risk loci (AD risk genes) from the Alzheimer’s Disease Sequencing Project (ADSP, https://adsp.niagads.org/), which were identified by a review of the literature by the Gene Verification Committee (GVC), pathway analysis, and by integration of genetic studies with myeloid genomics (Supplementary Table1-2). We applied our association approach to each of those genes by using the pretrained genomic LLM model to transform the personal DNA sequence from the gene-centric region to a diverse set of genomic and epigenomic features across different cellular contexts. Then a machine learning method (e.g., gradient boosting tree^37^ or support vector machine^38^) was used to causally associate the genomic LLM features to each imaging phenotype at a specific ROI and time point (see Methods). It is known that AD usually first damages the connections among neurons in parts of the brain involved in memory, especially the entorhinal cortex (EC) and hippocampus (HC) and then later affect other ROIs in the human brain^39,40^. It is seen that our association approach achieved a higher average correlation both across all ROIs and only two ROIs (EC and HC) compared to the genotype-based method for 12 out of 16 AD causal genes and 39 out of 56 AD risk gene (Figure 2A and Supplementary Figure 1). Our genomic LLM association approach ranked the most well-known AD causal gene APOE as the top gene, while the genotype-based method ranked it third. The average Pearson’s correlation coefficient (PCC) was improved by 29.7% using genomic LLM compared to the genotype-based method.

**Figure. 2.**
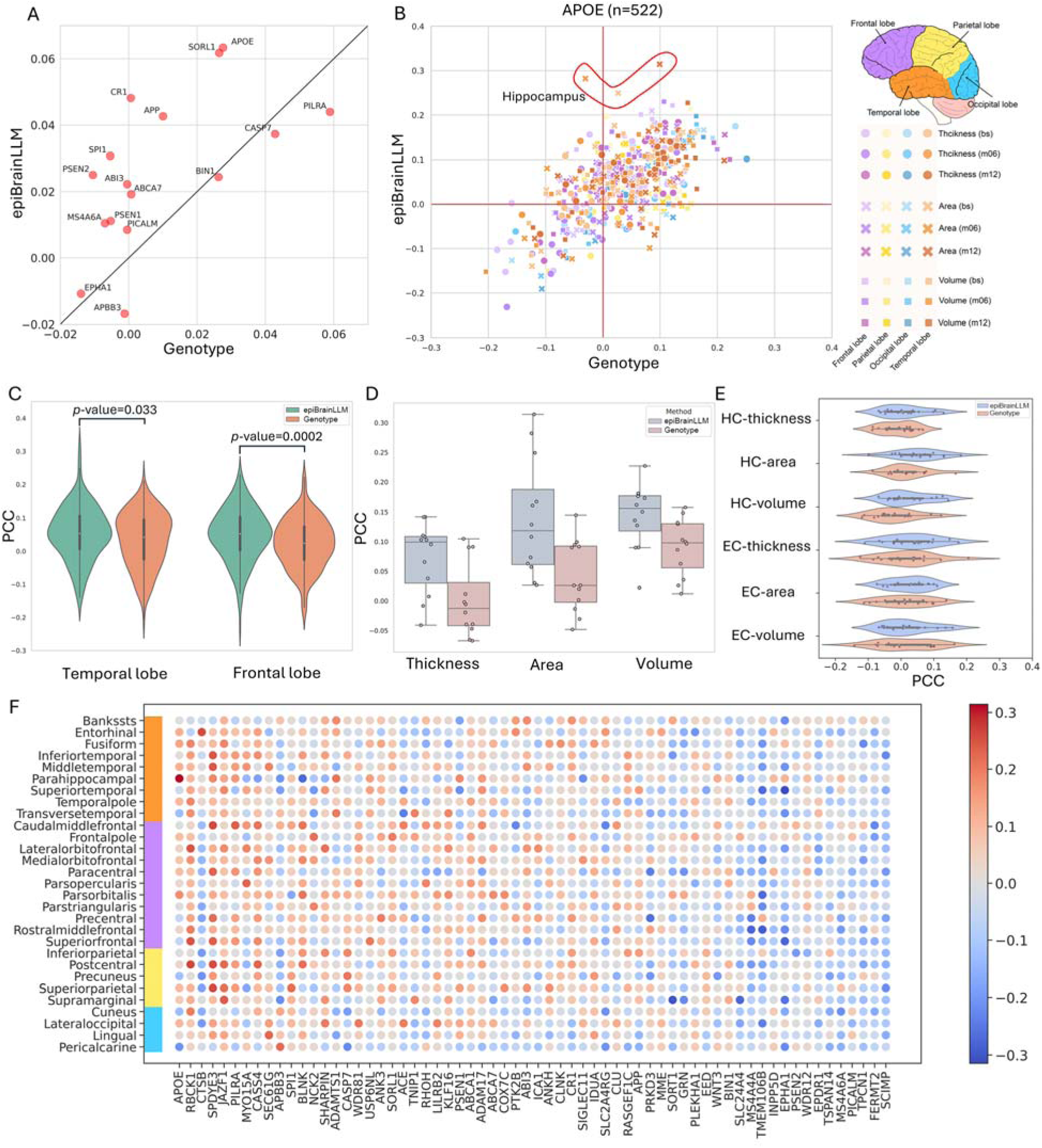
Gene-brain ROI causal relationship study. (A) epiBrainLLM achieves higher average Pearson’s correlation across all ROIs than the traditional genotype-based association approach in 12 out of 16 AD causal genes. (B) The association strength identified by epiBrainLLM and the genotype-based approach between APOE and 29 different human brain ROI across three imaging phenotypes and three time points. The brain surface area of hippocampus ranks the top in our approach. (C) Associations comparison of ROIs from temporal and frontal lobes for APOE gene. (D) Associations comparison of the two most important ROIs: HC and EC for APOE. (E) Detailed comparison across different imaging features for APOE. (F) Causal relationship map between AD causal/risk genes and brain ROIs where the genes were sorted by the maximum

Next, we took the most well-studied AD causal gene, APOE, for a detailed case study. By associating APOE with each brain ROI for different imaging trait and time point, hippocampus (HC)-area across three time points ranked the top 1-3 by our genomic LLM association approach while the genotype-based approach demonstrated even negative Pearson’s correlation (Figure 2B). We further analyzed the association strengths for brain ROIs from different lobes. The genomic LLM association approach illustrated a higher average Pearson’s correlation in temporal and frontal lobes, which are known to be highly impacted by the progression of AD (Figure 2C, *p*-value<0.05). When we further narrowed down the association analysis to the two most important AD-affected ROIs affected by AD, EC and HC, our genomic LLM approach outperformed the genotype-based method by a large margin by increasing the average PCC (across HC and EC) by 6.23%, 10.41%, 4.42% for three different imaging traits (thickness, area, volume), respectively (Figure 2D-E). Finally, to provide a holistic landscape of the association map between different genes and different brain ROIs, we displayed the causal association heatmap from epiBrainLLM by grouping different brain ROIs by brain lobes and ranked the genes according to the highest PCC (Figure 2F). Notably, APOE again ranked the first among all the AD causal and risk genes, with the HC ROI demonstrating the strongest association. These results suggest that the epiBrainLLM approach can provide stronger association signals to disease causal genes than the standard genotype-based approach.

### Improving AD risk assessment with epiBrainLLM

There is evidence showing that the progression of AD affects both the structure and function of the human brain^41^. MRI imaging features can be used to predict the risk of AD and contribute to AD diagnosis^42^. In our analysis, the three different imaging traits contribute differently to the AD status prediction while combining different imaging traits together could further improve the AD risk prediction by achieving an auROC of 0.720 (Figure 3A). When imaging data are not available, especially in the early stages, it is clinically useful to estimate the AD risk based on the genotype information. For example, genotype information of a 200kbp genomic region centered at the transcription start site (TSS) of APOE could lead to an auROC of 0.660 for predicting the AD risk while using the genomic LLM features for the same genomic region further increased the auROC to 0.701 (Figure 3C).

**Figure. 3.**
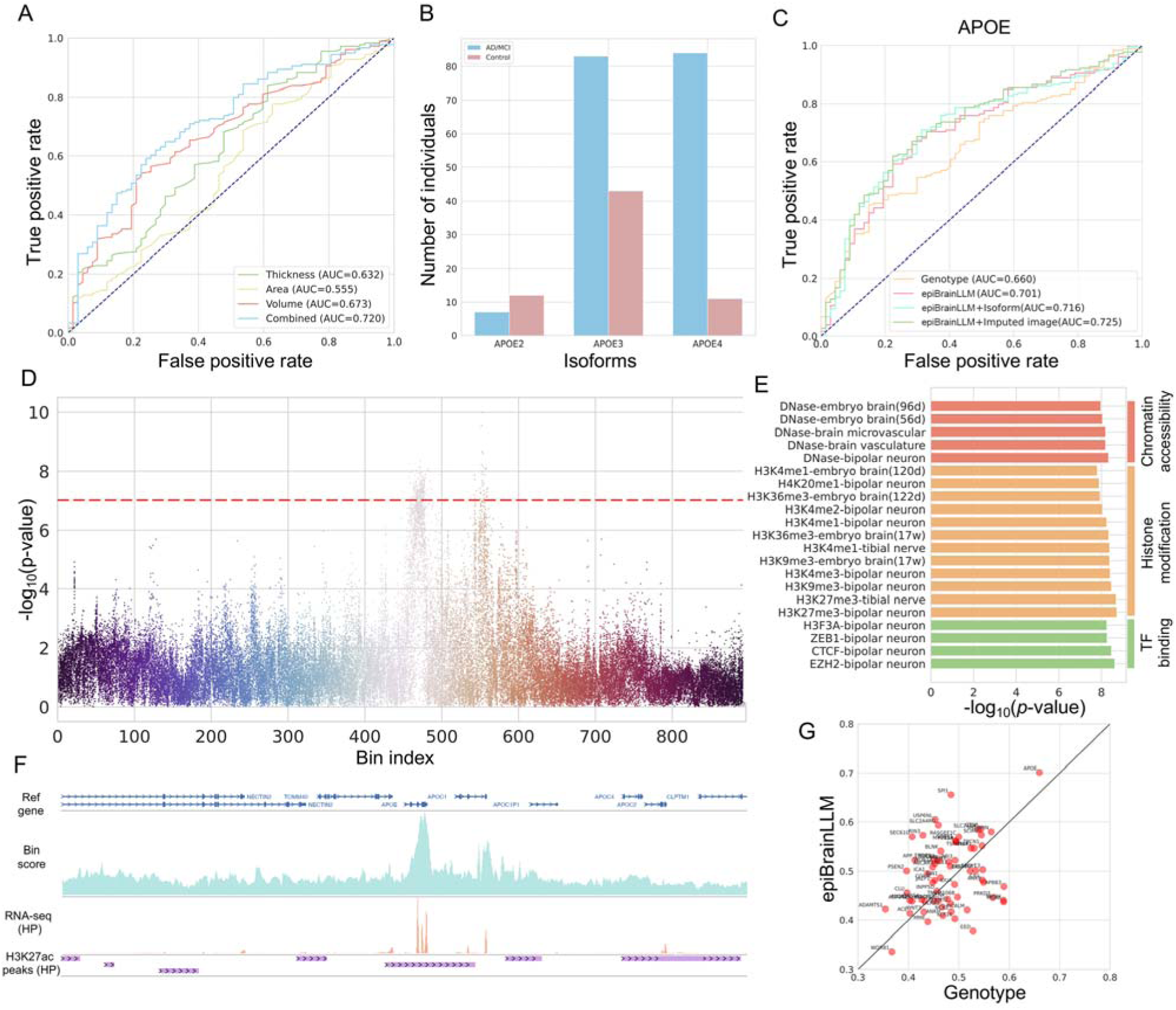
Causally Associating genes to AD disease status. (A) AD disease risk prediction with imaging features at baseline timepoint. (B) APOE isoforms in AD cases and controls. (C) AD disease risk prediction using genotype only, epiBrainLLM, epiBrainLLM plus gene isoform information, and epiBrainLLM plus imputed imaging features. (D) Association between genomic LLM features and AD disease. A *p*-value threshold 10^-7^ is used. (E) Top-enriched genomic LLM features, including chromatin accessibility, histone modification, and TF binding across different cellular contexts. (F) Annotations for the APOE-centric genomic region, including bin-level importance score (upper), RNA-seq of a HP brain tissue from ENCODE (middle), and H3K27ac peaks of a HP brain tissue from ENCODE (lower). (G) The gene-wise performance comparison of epiBrainLLM and the traditional genotype-based method in AD risk prediction. Our approach outperformed the genotype-based approach in 40 out of 64 (62.5%) AD causal/risk genes.

Next, we explored two directions to further improve the AD risk prediction with genomic LLM. First, we observed that several major APOE isoforms have significantly different distribution in AD cases and controls (Figure 3B). By combining the APOE isoforms information together with genomic LLM features, we further improved the AD risk prediction from an auROC of 0.701 to 0.716 (Figure 3C). Gene isoforms reveal the properties of proteins at the DNA level, complementing the genomic LLM features. Second, we investigated how to use the imputed imaging features (e.g., imaging features predicted from the genomic LLM features) to help improve the AD risk prediction. Using the imputed imaging features alone only yielded an auROC of 0.557 (Supplementary Figure 2). Combining the imputed imaging features with genomic LLM features further improved the AD risk prediction performance by achieving an auROC of 0.725 (Figure 3C, see Methods), which is comparable to the performance of using real imaging features (0.720). These results suggest a promising direction for improving AD risk prediction by leveraging the power of genomic LLM while the genotype data is available, but imaging data is not.

Additionally, we performed genomic LLM features association study by examining the distribution difference of each genomic LLM feature in case and control groups (see Methods). 286 genomic LLM features from 44 bins covering 5632 bps in the APOE region were identified by setting a *p*-value threshold of 10^-7^ (Figure 3D). The top-enriched genomic LLM features include chromatin accessibility, histone modification, and TF binding across various cellular contexts (Figure 3E). We noted that “bipolar neuron” is a highly enriched cellular context, and the current AD research has not extensively detailed the specific involvement of bipolar neurons, which are primarily found in sensory systems^43^. Existing studies showed that AD markers Aβ and tau pathology can spread to various regions, including those involved in sensory processing^44^. Our findings suggest a potential indirect relationship between bipolar neurons and AD disease that requires further exploration.

After analyzing each of the genomic LLM features across different bins, we also tried to identify the most important bins contributing to AD risk. We combined *p*-values for genomic LLM features within each bin to obtain the bin-level importance score (see Methods). Bins around the APOE and APOC1 genes contributed the most to the AD risk (Figure 3F). Multiple studies have shown that APOC1 polymorphisms may be associated with AD risk, supporting our findings^45,46^. We further displayed the RNA-seq and H3K27ac peaks of a human brain hippocampus (HC) tissue from the ENCODE database^47^ for the same genomic regions, showing consistency with the bin scores. Finally, we conducted systematic experiments to determine whether the genomic LLM could improve AD risk prediction based on the AD causal/risk genes. We combined the 16 AD causal genes and 56 AD risk genes and obtained a unique list of 64 AD-related genes. The genomic LLM-based approach outperforms the genotype-based approach in 62.5% of the genes while APOE again ranked the first for both methods (Figure 3G). If we combined the 64 AD-related genes together, our approach achieved an auROC of 0.731, compared to 0.698 of the genotype-based method (Supplementary Figure 3, see Methods).

### Facilitating longitudinal analysis of AD risk with epiBrainLLM

Longitudinal studies have been extensively conducted to characterize the structural changes of human brain over time, enhancing our understanding of AD disease progression at different stages^48^. In this study, we focused on longitudinal analysis in order to help classify the stable MCI (sMCI) versus progressive MCI (pMCI) where the pMCI patients eventually develop Alzheimer’s disease (AD) or dementia.

We first examined whether the prediction performance of imaging traits varies across different timepoints. We showed PCC of the 16 AD causal genes in the two most important AD-related brain ROIs (HC and EC) across three time points (Figure 4A). For further investigation, we also showed the PCC for APOE gene across time for different imaging traits (Figure 4B). Next, we explored how to leverage genomic LLM to distinguish sMCI from pMCI using the imaging features at early time point (e.g., baseline). The progressive trajectories of the 246 patients showed different transition patterns (Figure 4C and Supplementary Figure 4). We noted that combining three different imaging traits together at the baseline time point demonstrated an auROC of 0.676 (Figure 4D). Using the genomic LLM features together with the imaging traits at baseline further improved the prediction performance to an auROC of 0.707 (see Methods). Additionally, we showed that genomic LLM consistently led to improvement of auROC by 7.5% using imputed imaging traits (Supplementary Figure 5). These results indicate that genomic LLM can assist the diagnosis of AD disease progression and transition.

**Figure. 4.**
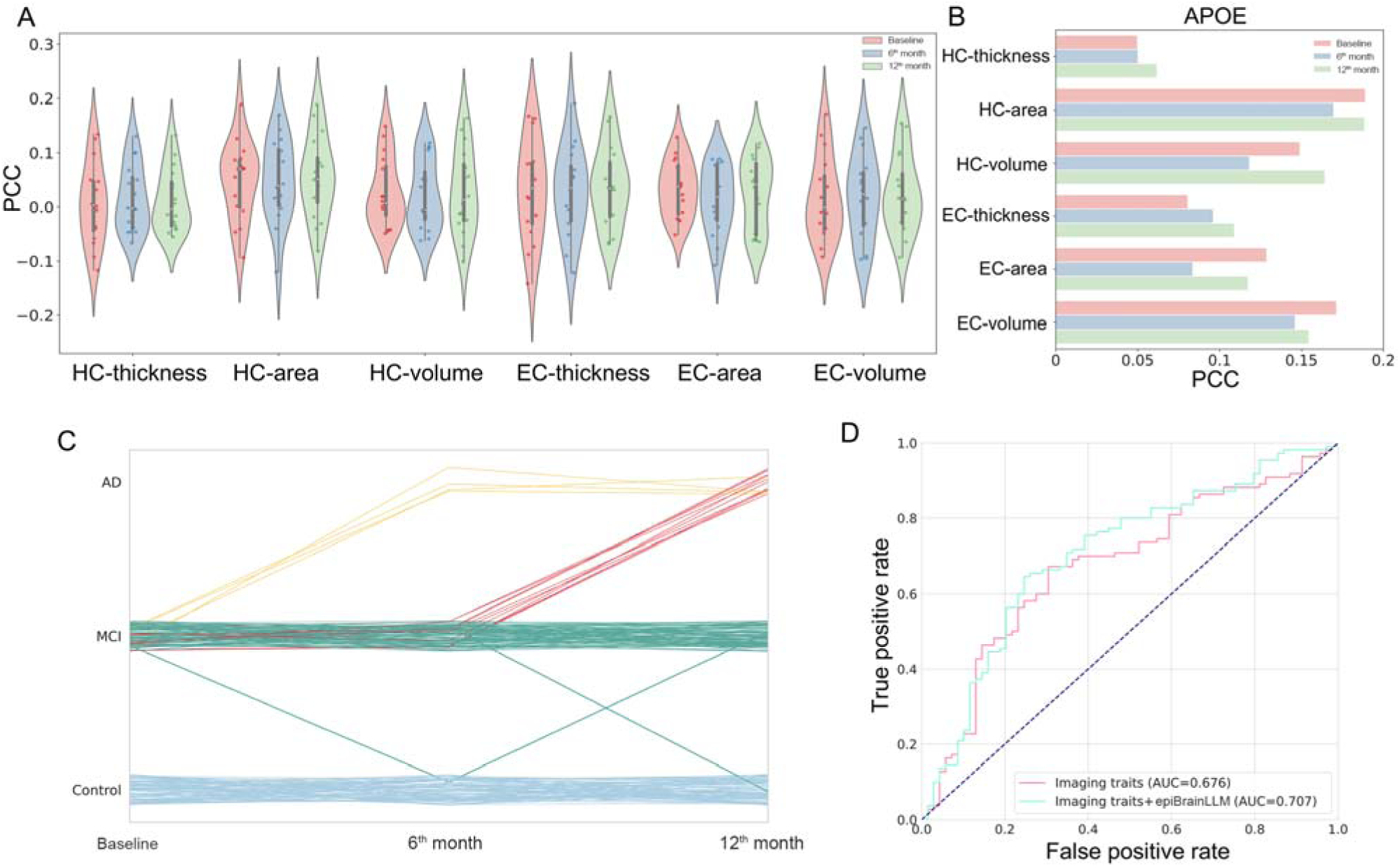
AD longitudinal analysis with epiBrainLLM. (A) The association between AD causal genes and HC, EC ROIs across three time points. (B) The association between APOE and HC, EC ROIs across three time points. (C) The AD disease trajectories of 246 participants in the first three time points. Different color represents different transition pattern. (D) Distinguishing stable MCI from progressive MCI with imaging traits at baseline time point, imaging traits plus genomic LLM features.

### Associating genotype to clinical phenotypes with epiBrainLLM

We further collected 785 individuals with both WGS and multiple types of clinical phenotype data, including amyloid beta (A*β*), total tau (t-tau), hyperphosphorylated tau (p-tau), mini mental state examination (MMSE), and functional activities questionnaire (FAQ). We used the similar two-stage epiBrainLLM approach to associate each of the AD causal or risk gene to a specific clinical phenotype. Note that we imputed a small fraction of missing values in the clinical phenotype data (Supplementary Figure 6). By comparing epiBrainLLM to the genotype-based approach, epiBrainLLM demonstrates a significantly higher association correlation in both biomarker phenotypes (A*β* and t-tau) and cognitive phenotypes (MMSE and FAQ) while the performance improvement is not significant on p-tau phenotype (Figure. 5A). APOE gene is again the most significant gene underlying the A*β* and FAQ phenotypes by both epiBrainLLM and genotype-based approach (Figure. 5C). Next, we explored whether the predicted clinical phenotype, such as a protein biomarker, is useful in predicting the AD status transition in the early stage. The 785 individuals were categorized into six different classes (Figure. 5B) where distinguishing the stable MCI (MCI to MCI, sMCI) from progressive MCI (MCI to AD, pMCI) is of particular importance for understanding disease progression and predicting conversion to AD. By integrating the information of all the 64 AD-related genes (see Methods), epiBrainLLM achieves an auROC of 0.665 in distinguish sMCI and pMCI patients (Figure. 5E). If we combined the genomic LLM features from multiple genes together with imputed biomarker expression (e.g., A*β*) at the baseline timepoint, the auROC is further improved from 0.665 to 0.683. Such results suggest that the imputed biomarker features from genomic LLM is beneficial to further improve the clinical

**Figure. 5.**
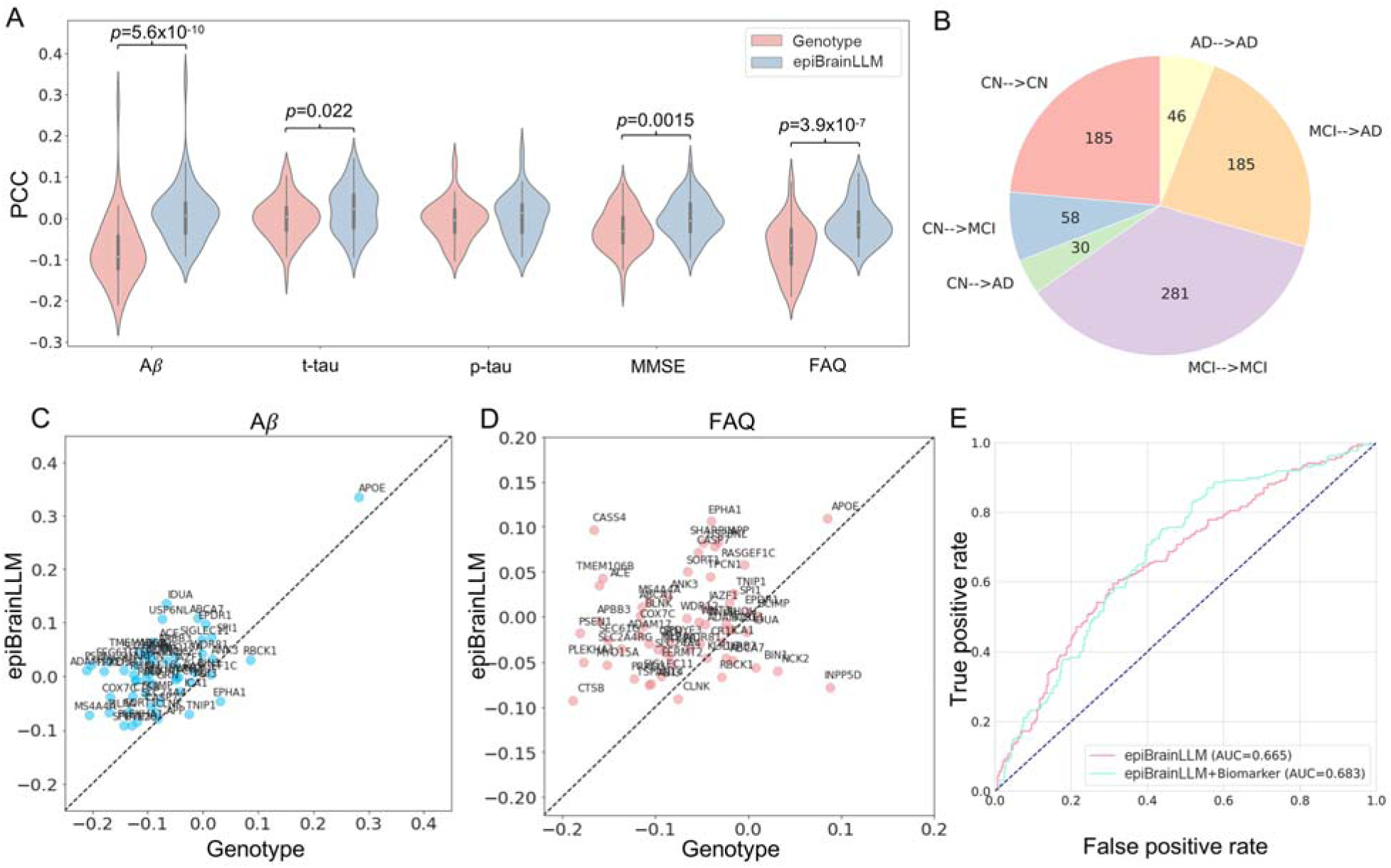
AD clinical phenotype association. (A) Comparison between epiBrainLLM and genotype-based approach on five different clinical phenotypes. (B) Six patterns of AD transition in 785 individuals. (C) Gene-wise association analysis between genotype and A*β* using epiBrainLLM and genotype-based approach. (D) Gene-wise association analysis between genotype and FAQ using epiBrainLLM and genotype-based approach. (E) Distinguishing sMCI from pMCI using epiBrainLLM by integrating information from multiple genes. The imputed biomarker information further improve the distinguishing power diagnosis of AD conversion.

## Discussion

In this study, we present a novel framework that leverages genomic large language models (LLMs) to enhance the association analysis between genetic variants and Alzheimer’s disease (AD) phenotype and imaging phenotypes. Our approach, which transforms personal DNA sequences into rich genomic and epigenomic features, outperformed traditional genotype-based method in associating AD-related genes with brain regions of interest (ROIs) and in predicting AD risk. Our two-stage approach epiBrainLLM can be seen as a more comprehensive extension of transcriptome-wide association studies (TWAS) as our approach considers multiple genomic and epigenomic signals as intermediate variables, not just gene expression. The diverse set of imputed intermediate features have the great potential in improving the causal discovery. In our analysis, we also identified previously unrecognized potential involvement of bipolar neurons in AD pathology through our genomic LLM features association study. Furthermore, our longitudinal analysis demonstrated the method’s utility in distinguishing between stable and progressive mild cognitive impairment (MCI).

Several extensions the model can further improve our approach. First, we showed that integration of multiple genes could improve the phenotype prediction compared to provided gene-wise association analysis. Further combining the genome-wide genomic LLM features from different linkage disequilibrium (LD) blocks may lead to novel discoveries. Second, integrating information from multiple genomic regions increases the dimensionality of the intermediate genomic and epigenomic features. Applying modern dimension reduction and causal inference tools^49,50^ could further help elucidate the causal mechanism underlying the complex diseases. Third, the integration of other LLMs in place of Enformer^24^ could offer additional improvements. For instance, models like EpiGePT^25^, designed to predict epigenomic signals given a specific cellular context, could help us better understand the genotype-phenotype associations in a context-specific manner.

While our study focused on known AD-related genes and a limited sample size, these findings suggest that integrating genomic LLMs with traditional genetic and imaging data offers a promising new direction for understanding the complex regulatory mechanisms underlying AD progression. Future work should validate these findings in larger, diverse cohorts and explore the application of this framework to other complex diseases. Overall, our study provides a novel perspective for interpreting genetic variation in AD and potentially paving a new way for identifying new biological features, prognostic markers, and therapeutic targets.

## Online Methods

### Whole genome sequencing data (WGS) preprocessing

The WGS data with .vcf format of 808 individuals were downloaded from Alzheimer’s Disease Neuroimaging Initiative (ANDI) database (https://adni.loni.usc.edu/). The WGS data were recalled using the BWA and GATK HaplotypeCaller pipeline^51^. We designed the following steps to construct the personal genome for each individual. 1) Variant Filtering: Our current framework focuses on single nucleotide variants (SNVs) only. We removed all the insertion and deletion (indels) from the .vcf file and only kept SNVs using vcftool^52^ (version 0.1.15). 2) Genotype Phasing: We employed the reference-free Beagle software^53^ (version 5.4) to phase the genotypes. This phasing step is crucial for differentiating between maternal and paternal alleles, thus playing an important role in the allele-specific analysis. 3) Personal Genome Construction: We utilized the vcf2diploid tool^54^ (version v0.2.6a) to reconstruct individual maternal and paternal genomes. This process generates two distinct haploid genome sequences for each subject, representing the respective maternal and paternal genetic contributions. After the above processing steps, each individual has a maternal and a paternal genome sequence containing single nucleotide genetic variants (e.g., SNPs).

### MRI imaging data preprocessing

The structural MRI (sMRI) imaging data (3D whole brain scans) in .nii format for 639 individuals were downloaded from the ADNI database with entry name “ADNI1:Complete 1Yr 1.5T”. Each individual has the sMRI imaging data of at least three time points, including baseline, 6^th^ month, and 12^th^ month. We employed the FreeSurfer software (version 7.3.2) “recon-all” command with default parameters to preprocess the sMRI data for each individual per time point. Note that FreeSurfer^55^ is able to handle multiple replicates if per individual and per time point contains multiple sMRI images. The Desikan-Killiany Atlas (Desikan 2006)^56^ is used for the cortical parcellation. After cortical parcellation, each hemisphere cortical surface of the human brain will be parcellated into multiple regions of interest (ROIs) (Supplementary Table 4). Each ROI is annotated with a unique neuroanatomical label based on probabilistic information estimated from a manually labeled training set by FreeSurfer. Three commonly used imaging phenotypes, including cortical thickness, cortical area and volume, are extracted for each individual at three different time points. A standard normalization is applied to the each of the imaging phenotype at each time point across individuals to by removing the mean and scaling to unit variance.

### Genomic large language model

A genomic large language model (LLM) is an advanced AI system designed to understand the complex relationship between genomic sequences and genomic signals derived from either functional annotations or Next-generation Sequencing (NGS) experiments. Leveraging the architecture and capabilities of transformer-based LLMs, the genomic LLMs are specifically trained on vast amounts of genomic sequences and related biological information. The core transformer architecture uses self-attention mechanisms to process input sequences, allowing it to consider the entire context of a sequence simultaneously, rather than locally or sequentially^57^. The inherent similarities between DNA sequence and natural language provide a promising perspective for using language models to decipher the functional implications of a genetic variants that occurs in the human genome. More details of applying transformer-based language model to bioinformatics can be found in a recent review paper^21^.

In our work, we chose the Enformer^24^, one of the state-of-the-art genomic language models in the field, which was trained based on the reference human genome (e.g., hg19) and a large panel of public genome-wide genomic and epigenomic data collected from ENCODE^47^, GEO^58^, FANTOM^59^ databases. Enformer aims at building a mapping relationship between the DNA sequence of a genomic region and multiple genomic and epigenomic signals of the corresponding region. Enformer takes a reference DNA sequence from a 196,608 bp genomic region consisting of 1536 bins (bin size: 128 bp) as input and employs a transformer encoder architecture to predict 5,313 human genomic and epigenomic signals for each of the central non-overlapping 896 bins given the input genomic region (320 bins in both sides of the input genomic region are cropped). The comprehensive output signals include gene expression, chromatin accessibility, histone modifications, TF binding profiles across a diverse set of cellular contexts (https://github.com/calico/basenji/blob/master/manuscripts/cross2020/targets_human.txt). By applying the pretrained Enformer model to a personal DNA sequence, the predicted personal genomic and epigenomic signals could reflect how the genetic variants contribute to the changes in various molecular phenotypes. The pretrained Enformer model was downloaded from https://www.kaggle.com/models/deepmind/enformer/TensorFlow2/enformer/1 and used in our study to convert the personal genetic variants into a diverse set of personal molecular phenotypes. Note that our framework is also highly flexible to incorporate new state-of-the-art genomic LLM models that may emerge in the future.

### Translation from genetic variants to genomic and epigenomic signals

In this study, we focus on extracting the genomic and epigenomic signals for a specific gene using a pretrained genomic LLM model (e.g., Enformer). We designed the following steps for a gene-wise feature extraction analysis given the personal WGS data. 1) We define a gene-centric genomic region centered at the transcription start site (TSS) of the gene. The length of genomic region is 196,608 bp for AD disease phenotype where the central non-overlapping 896 bins (bin size: 128 bp) are considered. For more complex and fine-grained imaging phenotypes, we choose a longer genomic region consisting of 425,984 bp where the central non-overlapping 2688 bins (896X3) are considered. Both the paternal and maternal DNA sequences of an individual from an Enformer input genomic region are extracted and represented in a one-hot encoding format. 2) We take the pretrained genomic LLM model (e.g., Enformer) as a feature extractor and apply it to both the paternal and maternal DNA sequences of an individual.

### Dimension reduction of genomic LLM features

Through the above strategy, the Enformer-based feature extractor could help generate a large number of genomic and epigenomic features, which yield approximately 9.5 million per individual (5,313 features per bin × 896 bins × 2 haploid genomes) per Enformer input genomic region. Such ultra-high dimensional features raise an urgent challenge in the follow-up phenotype prediction tasks to handle a vast number of predictor variables. We proposed the following strategies, including feature selection and local principal component analysis (PCA), and haploid aggregation to mitigate this high dimensionality issue. 1) Feature Selection. we select a subset of the AD-related signals out of the 5,313 genomic and epigenomic signals. Specifically, we selected 77 signals that are related to brain, neuron or nerve, which could significantly reduce the genomic LLM feature dimension by 98.5% (Supplementary Table 3). This feature selection could reduce the individual-level genomic LLM features from 9.5 million to 128k for each Enformer input region, which is still high-dimensional compared to the small sample size. Note that this step is only applicable to AD disease phenotype. 2) Local PCA. We further apply a local PCA to each bin-wise features across individuals where we only keep the top-k PC components in order to capture the majority of the cross-individual variation in the original high-dimensional data. We set k=4 for AD disease phenotype and k=7 for imaging phenotypes. The local PCA can further reduce the dimension of the genomic LLM features, which leads to a much more manageable scale for the follow-up phenotype prediction tasks. 3) Haploid Aggregation. Since we extracted the genomic LLM features for both paternal and maternal genome sequence, which indicate the parent of origin, we need to further aggregate the paternal features and maternal features together to finally get the individual-level genomic LLM features. Mean aggregation is used as default for AD disease phenotype. Note that we choose to concatenate the paternal and maternal features for imaging phenotypes. The above proposed dimension reduction strategies are crucial for us to get a manageable scale and informative representation of the original high-dimensional genomic LLM features and make the subsequent phenotype prediction tasks statistically robust and computationally feasible.

### Phenotype prediction

After obtaining the genomic LLM features from the personal genome sequence around a gene, we can then apply any type of machine learning algorithm to perform the phenotype prediction tasks. Since the genomic LLM feature extractor is a highly non-linear transformation, we provide several non-linear machine learning methods, including support vector machine (SVM)^38^ with a nonlinear radial basis function (RBF) kernel and gradient boosting decision trees (GBDT)^37^. SVM is implemented with the Sklearn library^60^ with default parameters. GBDT is also implemented with the Sklearn library where 40 trees are used. We use SVM for imaging phenotypes prediction and GBDT for AD clinical phenotype prediction. To systematically evaluate the phenotype prediction performance, five-fold cross-validation is used to avoid over-fitting.

The genotype-based approach directly uses the genotype information of SNPs (0, 1 or 2) as input for the machine learning method while our genomic LLM approach takes the genomic LLM features as input for the same machine learning method. We employ a fair comparison setting to demonstrate the performance of our method. For example, our epiBrainLLM approach only uses the features from the central 114,688 bp (896 bins) out of the whole 196,608 bp (1536 bins) input genomic region for AD disease phenotype. We use all the SNPs within the 196,608 bp input genomic region for the genotype-based method.

In order to combine the information of multiple genes, we propose a meta learning approach where the prediction scores of either our approach or genotype-based method for each gene are concatenated and fed to an additional GBDT model. Note that the prediction score is one-dimensional in AD status prediction and six-dimensional in AD transition prediction as there are six transition patterns. This allows users to integrate the information from multiple genes in order to get a better clinical phenotype prediction performance.

### Imaging features imputation

To get imputed imaging features at a specific time point (e.g., baseline), we used the fitted machine learning model (still in a five-fold cross-validation setting) in last subsection to predict the three different types of imaging traits of 29 brain ROIs. The predicted features of different imaging traits were concatenated for further prediction.

### AD clinical phenotype imputation

The five different AD clinical phenotypes (A*β*, t-tau, p-tau, MMSE, and FAQ) contains different level of data missing with missing rate ranging from 0% to 24.08%. We used multiple imputation by chained equation (MICE) algorithm^61^ implemented by sklearn library^60^ with default parameters for data imputation.

### Genomic LLM features enrichment

To evaluate whether a genomic LLM feature has different distribution in AD case and control groups. We perform large-scale statistical hypothesis tests for all the genomic LLM features. Since the genomic LLM features are continues, a two-sample t-test is used to test the significance and calculate the *p*-value for each genomic LLM feature. Since genomic LLM features are allele-specific, the Fisher’s method is used to combine paternal and maternal *p*-values.

In order to get the bin-level importance score, we also adopt the idea of Fisher’s method by calculating the statistic −2 ∑*_i_* log (*p_i_*) where i indexes across features within a bin. The importance score is then obtained by dividing the above statistic by a normalizing constant, which is maximum value among all the bins.

### Evaluation metrics

When the phenotype is binary (e.g., whether the individual is an AD patient or not), the commonly used the area under the receiver operating characteristic curve (auROC) is used for evaluating the prediction performance. When the phenotype is continuous (e.g., cortical thickness), the Pearson correlation coefficient (PCC) is used for evaluating the prediction performance.

## Data availability

The raw data in this study can be accessed through Alzheimer’s Disease Neuroimaging Initiative: ADNI (https://adni.loni.usc.edu/). WGS data using BWA & GATK HaplotypeCaller pipeline were downloaded in the “Genetic Data” category. sMRI images were download in the “Image Collections” category under the “ADNI1:Complete 1Yr 1.5T” entry. The annotation data, including gene expression and histone modification data, were originally from ENCODE database and visualized with WashU Epigenome Browser (https://epigenomegateway.wustl.edu/).

## Code availability

Code is publicly available at https://github.com/SUwonglab/AD-genomicLLM.

## Acknowledgement

The works of Q.L. was partially supported by the National Human Genome Research Institute (NHGRI) of the National Institutes of Health (NIH) under Award Number K99HG013661. The works of Q.L., W.Z., and W.H.W were partially supported by NIH grants R01HG010359 and R01HG007735. The work of L.L. was supported by NSF grant CIF-2102227, and NIH grants R01AG061303 and R01AG062542. The work of H.Z. was partially supported by the National Institute On Aging (NIA) of the NIH [U01AG079847, 1R01AG085581, RF1AG082938]. Data used in preparation of this article were obtained from the Alzheimer’s Disease Neuroimaging Initiative (ADNI) database (adni.loni.usc.edu). As such, the investigators within the ADNI contributed to the design and implementation of ADNI and/or provided data but did not participate in analysis or writing of this report. A complete listing of ADNI investigators can be found at: http://adni.loni.usc.edu/wp-content/uploads/how_to_apply/ADNI_Acknowledgement_List.pdf.

