## Supplementary Materials for "Leveraging genomic large language models to enhance causal genotype-brain-clinical pathways in Alzheimer’s disease"

[Supplementary Figures](#_Toc493166363)	3

[Supplementary Tables](#_Toc493166364)	8

**Supplementary Figures**

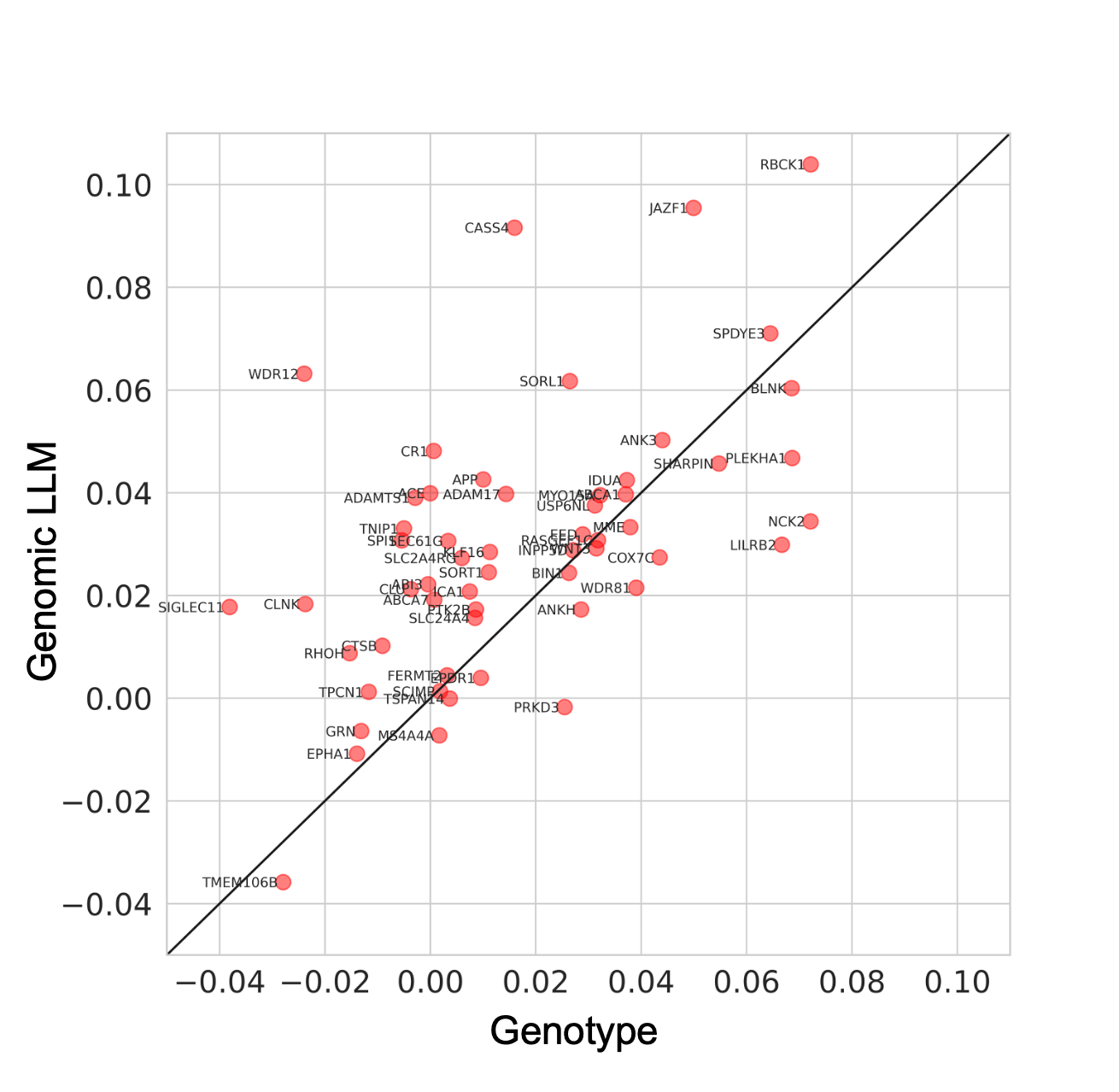

**Supplementary Figure 1.** The comparison of genomic LLM-based approach and the genotype-based approach on associating AD risk genes to human brain ROIs. Each dot represents an AD risk gene and the average Pearson’s correlation coefficient (PCC) is shown. The genomic LLM-based approach outperformed the genotype-based approach in 39 out of 56 AD risk genes.

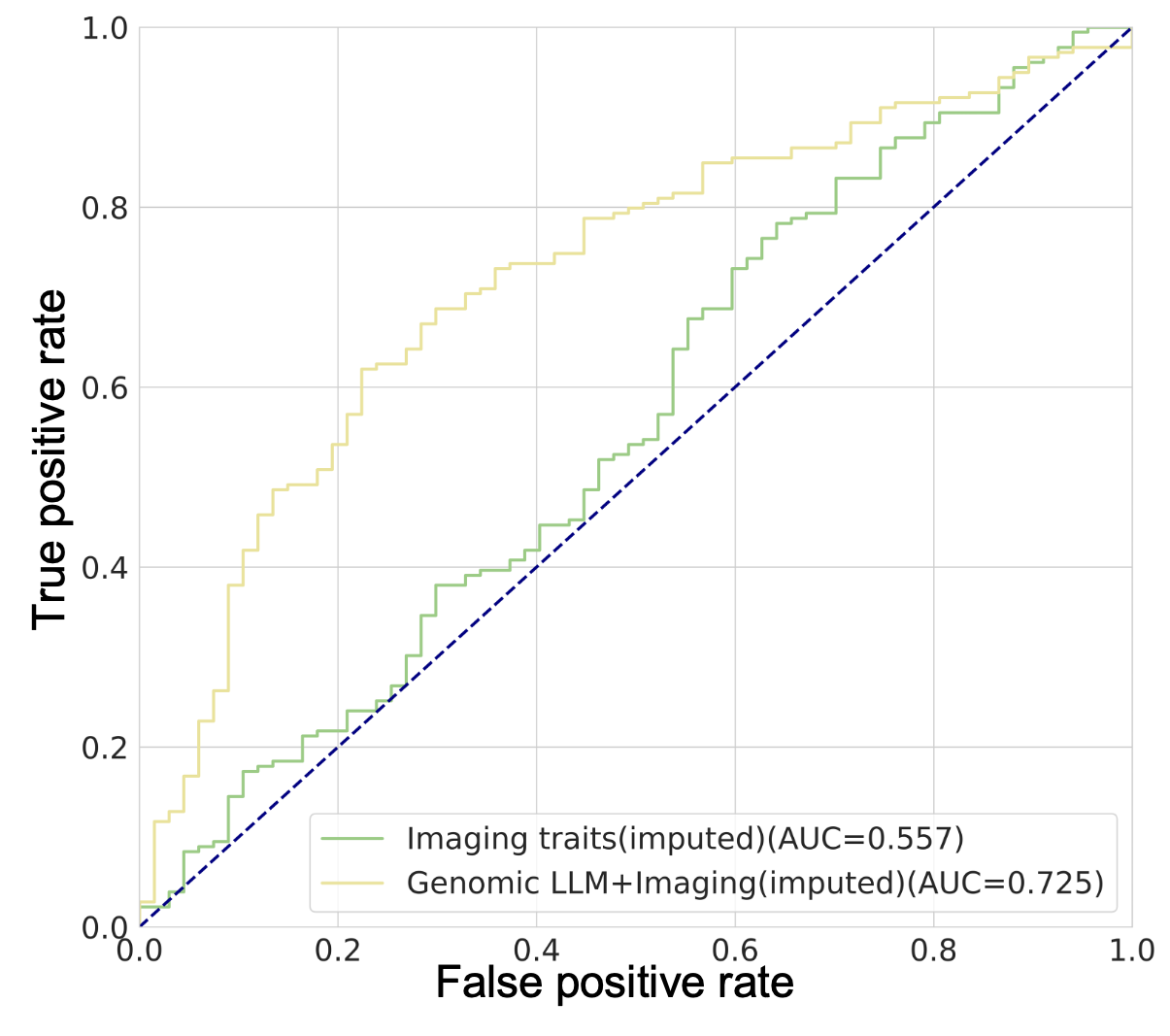

**Supplementary Figure 2.** AD disease risk prediction with imputed imaging features (green) and combining genomic LLM features together with imputed imaging features (yellow).

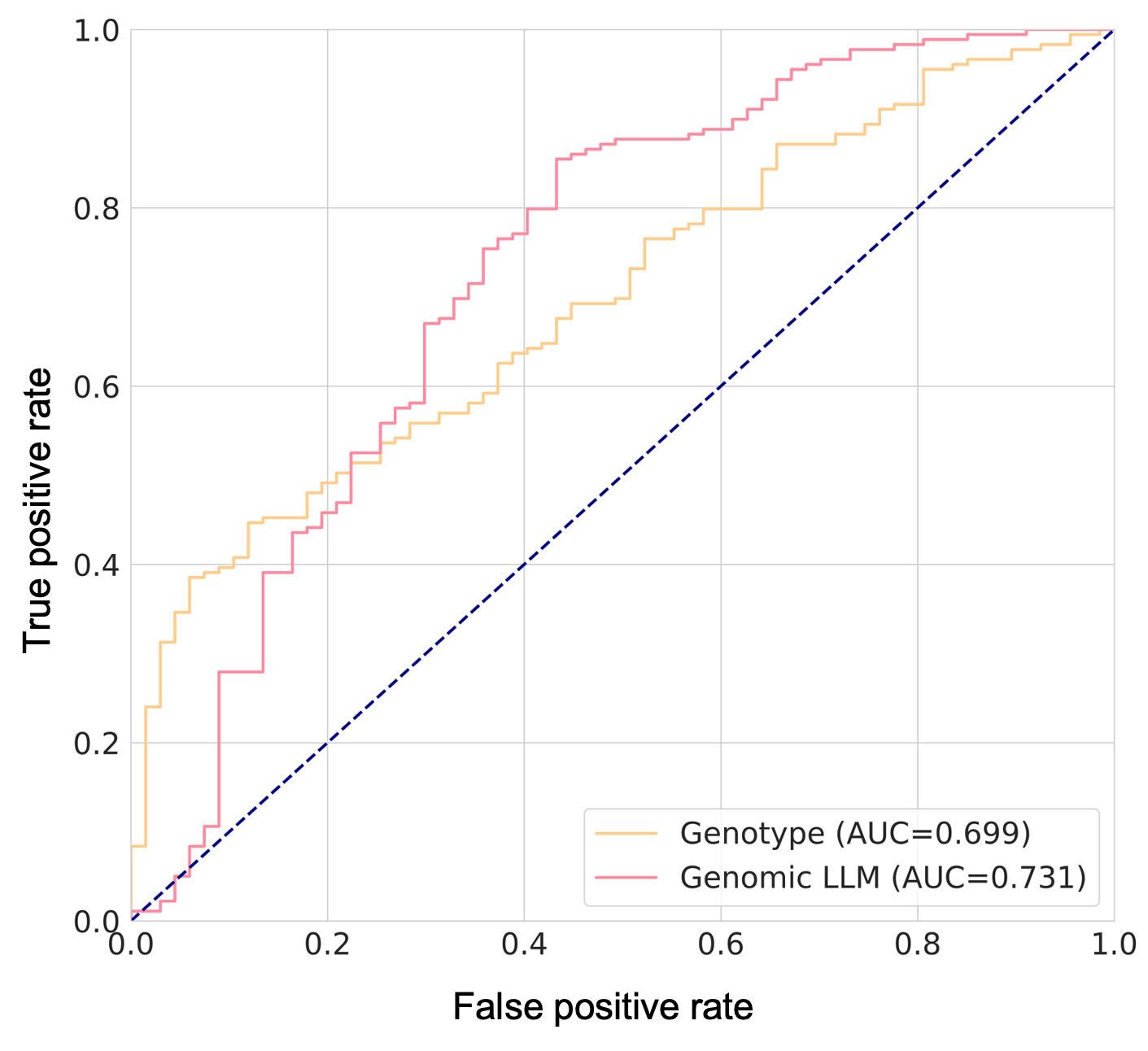

**Supplementary Figure 3.** Combined information of 64 AD-related genes to predict AD status. Our approach achieves an auROC of 0.731 while the genotype-based method achieves an auROC of 0.699

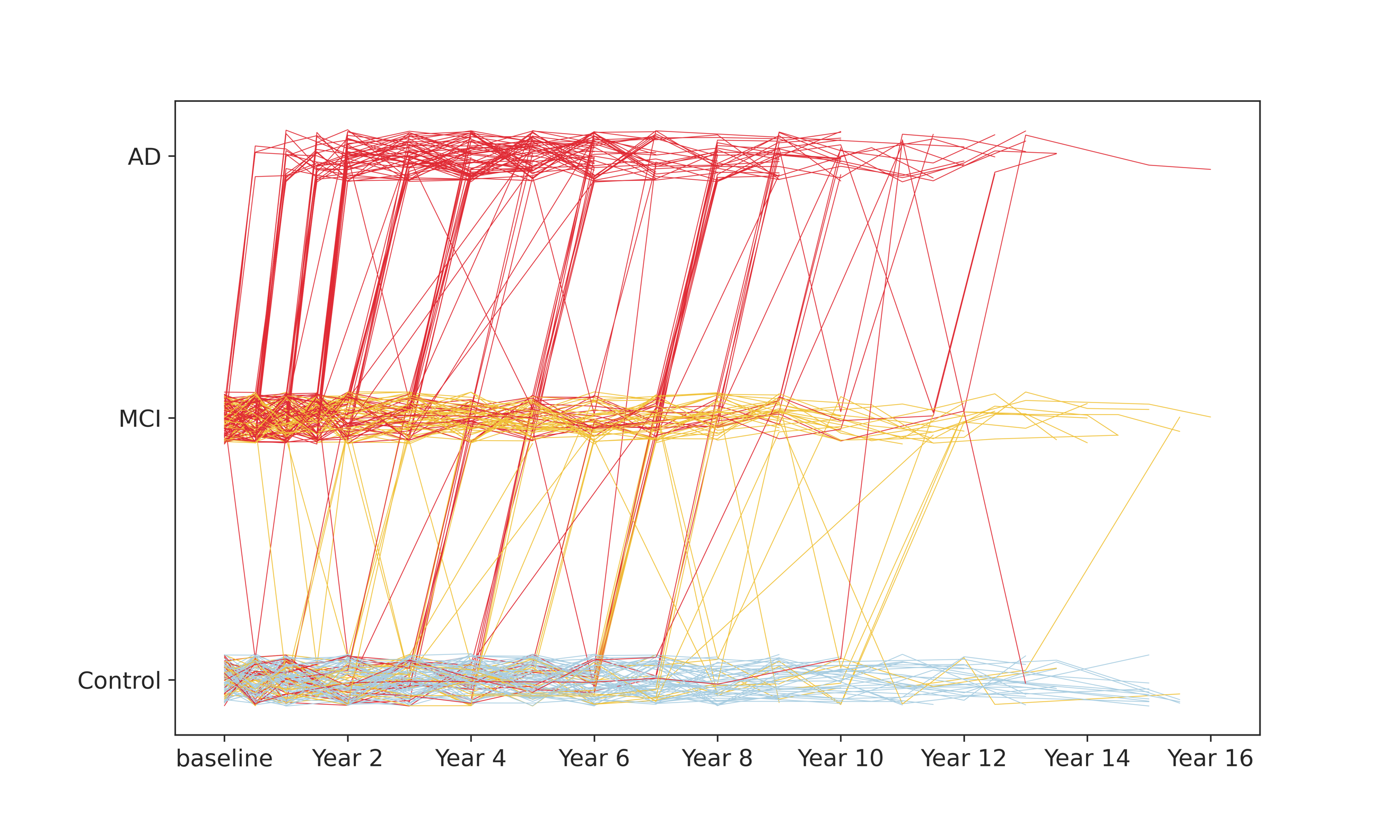

**Supplementary Figure 4.** AD trajectories of 246 patients in our study up to 16 years based on the records from ADNI database. Color represents final clinical AD diagnosis where blue for CN, yellow for MCI, and red for AD.

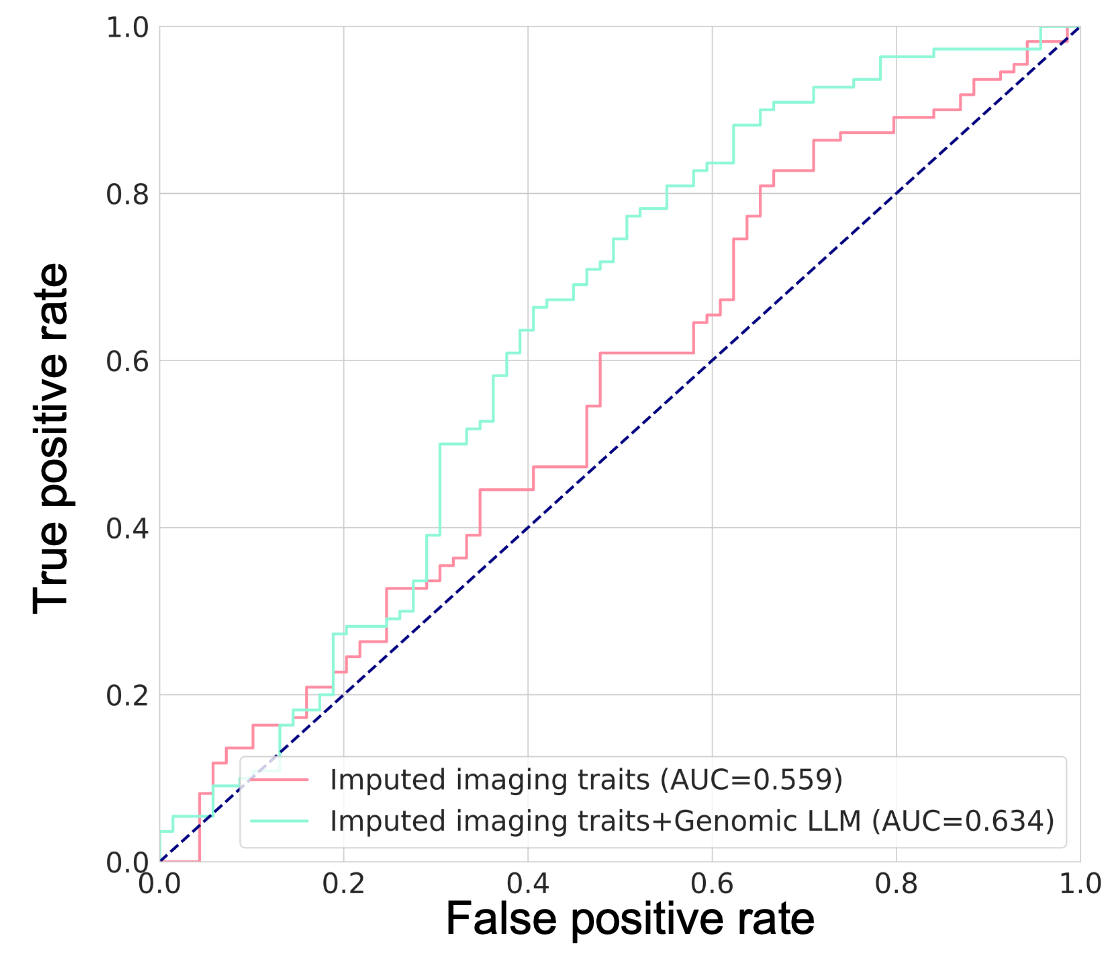

**Supplementary Figure 5.** AD disease risk prediction using imputed imaging features (red) and using imputed imaging features combined with genomic LLM features (green).

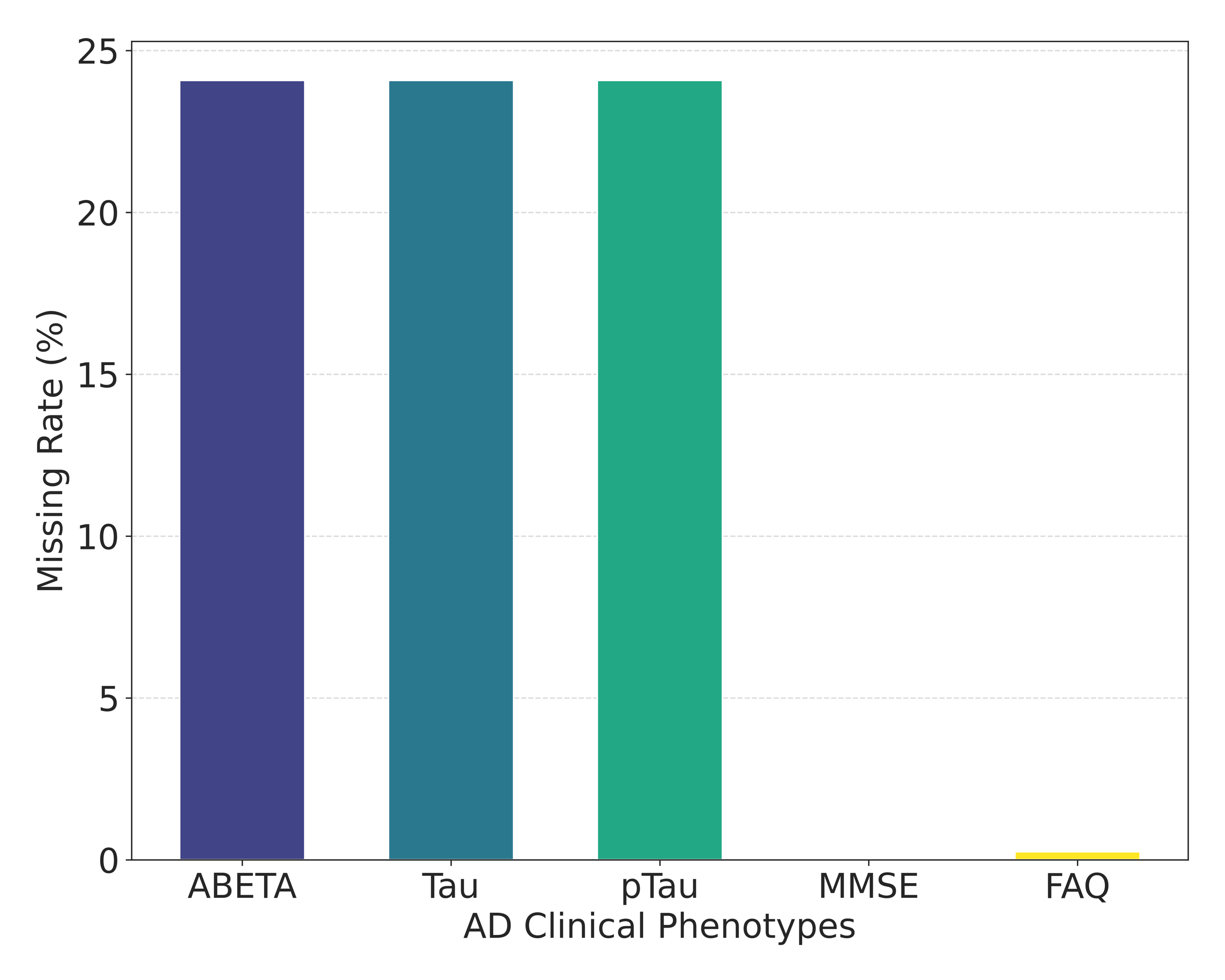

**Supplementary Figure 6.** Missing rate for different type of AD clinical phenotype.

**Supplementary Tables**

**Supplementary Table 1**. List of Alzheimer's disease 16 causal genes from Alzheimer’s Disease Sequencing Project (ADSP). Genes from chr6, chr15, chr16 from the original list were excluded as the WGS processing step was not finished within the time limit (7 days).

| Gene name | Chromosome | TSS position (hg19) | Strand |
| --- | --- | --- | --- |
| CR1 | chr1 | 207669472 | + |
| PSEN2 | chr1 | 227058272 | + |
| BIN1 | chr2 | 127864903 | - |
| APBB3 | chr5 | 139944189 | - |
| EPHA1 | chr7 | 143105985 | - |
| PILRA | chr7 | 99971067 | + |
| CASP7 | chr10 | 115438920 | + |
| MS4A6A | chr11 | 59952139 | - |
| SORL1 | chr11 | 121322911 | + |
| SPI1 | chr11 | 47400127 | - |
| PICALM | chr11 | 85780139 | - |
| PSEN1 | chr14 | 73603142 | + |
| ABI3 | chr17 | 47287588 | + |
| ABCA7 | chr19 | 1040101 | + |
| APOE | chr19 | 45409038 | + |
| APP | chr21 | 27543138 | - |

**Supplementary Table 2**. List of Alzheimer's disease 16 risk genes from Alzheimer’s Disease Sequencing Project (ADSP). Note that the “risk genes” are defined as the nearest gene of AD risk loci. Genes from chr6, chr15, chr16 from the original list were excluded as the WGS processing step was not finished within the time limit (7 days).

| Gene name | Chromosome | TSS position (hg19) | SNV | Strand |
| --- | --- | --- | --- | --- |
| SORT1 | chr1 | 109935979 | rs141749679 | - |
| CR1 | chr1 | 207669472 | rs679515 | + |
| ADAM17 | chr2 | 9695917 | rs72777026 | - |
| PRKD3 | chr2 | 37544222 | rs17020490 | - |
| NCK2 | chr2 | 106361519 | rs143080277 | + |
| BIN1 | chr2 | 127864903 | rs6733839 | - |
| WDR12 | chr2 | 203776949 | rs139643391 | - |
| INPP5D | chr2 | 233925035 | rs10933431 | + |
| MME | chr3 | 154797704 | rs16824536 | + |
| IDUA | chr4 | 980784 | rs3822030 | + |
| CLNK | chr4 | 10686386 | rs6846529 | - |
| RHOH | chr4 | 40198526 | rs2245466 | + |
| ANKH | chr5 | 14871887 | rs112403360 | - |
| COX7C | chr5 | 85913783 | rs62374257 | + |
| TNIP1 | chr5 | 150444692 | rs871269 | - |
| RASGEF1C | chr5 | 179636130 | rs113706587 | - |
| ICA1 | chr7 | 8301682 | rs10952097 | - |
| TMEM106B | chr7 | 12250847 | rs13237518 | + |
| JAZF1 | chr7 | 28220437 | rs1160871 | - |
| EPDR1 | chr7 | 37960920 | rs6966331 | + |
| SEC61G | chr7 | 54826939 | rs76928645 | - |
| SPDYE3 | chr7 | 99905324 | rs7384878 | + |
| EPHA1 | chr7 | 143105985 | rs11771145 | - |
| CTSB | chr8 | 11725646 | rs1065712 | - |
| PTK2B | chr8 | 27182995 | rs73223431 | + |
| CLU | chr8 | 27469268 | rs11787077 | - |
| SHARPIN | chr8 | 145159140 | rs34173062 | - |
| ABCA1 | chr9 | 107690527 | rs1800978 | - |
| USP6NL | chr10 | 11653679 | rs7912495 | - |
| ANK3 | chr10 | 62493284 | rs7068231 | - |
| TSPAN14 | chr10 | 82214037 | rs6586028 | + |
| BLNK | chr10 | 98031273 | rs6584063 | - |
| PLEKHA1 | chr10 | 124134093 | rs7908662 | + |
| SPI1 | chr11 | 47400127 | rs10437655 | - |
| MS4A4A | chr11 | 60048013 | rs1582763 | + |
| EED | chr11 | 85955805 | rs3851179 | + |
| SORL1 | chr11 | 121322911 | rs11218343 | + |
| TPCN1 | chr12 | 113659259 | rs6489896 | + |
| FERMT2 | chr14 | 53417815 | rs17125924 | - |
| SLC24A4 | chr14 | 92788924 | rs7401792 | + |
| WDR81 | chr17 | 1619816 | rs35048651 | + |
| SCIMP | chr17 | 5138083 | rs7225151 | - |
| MYO15A | chr17 | 18012019 | rs2242595 | + |
| GRN | chr17 | 42422490 | rs5848 | + |
| WNT3 | chr17 | 44896082 | rs199515 | - |
| ABI3 | chr17 | 47287588 | rs616338 | + |
| ACE | chr17 | 61562177 | rs4277405 | + |
| ABCA7 | chr19 | 1040101 | rs12151021 | + |
| KLF16 | chr19 | 1863564 | rs149080927 | - |
| SIGLEC11 | chr19 | 50464429 | rs9304690 | - |
| LILRB2 | chr19 | 54785033 | rs587709 | - |
| RBCK1 | chr20 | 388708 | rs1358782 | + |
| CASS4 | chr20 | 54987313 | rs6014724 | + |
| SLC2A4RG | chr20 | 62371210 | rs6742 | + |
| APP | chr21 | 27543138 | rs2154481 | - |
| ADAMTS1 | chr21 | 28217728 | rs2830489 | - |

**Supplementary Table 3**. The 77 selected genomic LLM features for AD disease out of 5513 Enformer features.

| Feature index | File ID | Experiment ID | Description |
| --- | --- | --- | --- |
| 485 | ENCFF399ISP | ENCSR626RVD | DNASE:bipolar neuron originated from GM23338 treated with 0.5 ug/mL doxycycline hyclate for 4 days |
| 1444 | ENCFF869ZLL | ENCSR000GGX | CHIP:H3K9me3:neuron originated from H9 |
| 1644 | ENCFF596ZSJ | ENCSR066BZZ | CHIP:CTCF:bipolar neuron originated from GM23338 treated with 0.5 ug/mL doxycycline hyclate for 4 days |
| 1998 | ENCFF043ACF | ENCSR175OYG | CHIP:H3K9me2:bipolar neuron originated from GM23338 treated with 0.5 ug/mL doxycycline hyclate for 4 days |
| 2223 | ENCFF380HMH | ENCSR247PYK | CHIP:H3K9me3:bipolar neuron originated from GM23338 treated with 0.5 ug/mL doxycycline hyclate for 4 days |
| 2375 | ENCFF011TOL | ENCSR301AEA | CHIP:H3K4me1:bipolar neuron originated from GM23338 treated with 0.5 ug/mL doxycycline hyclate for 4 days |
| 2409 | ENCFF548EWI | ENCSR314WYC | CHIP:H3K4me3:neuron originated from H9 |
| 2592 | ENCFF643URD | ENCSR375IXS | CHIP:H3K36me3:bipolar neuron originated from GM23338 treated with 0.5 ug/mL doxycycline hyclate for 4 days |
| 2721 | ENCFF286MEX | ENCSR418KUS | CHIP:ZEB1:bipolar neuron originated from GM23338 treated with 0.5 ug/mL doxycycline hyclate for 4 days |
| 2896 | ENCFF155KWT | ENCSR472SEY | CHIP:H3K27me3:bipolar neuron originated from GM23338 treated with 0.5 ug/mL doxycycline hyclate for 4 days |
| 2938 | ENCFF324STM | ENCSR484PTR | CHIP:H3K79me2:bipolar neuron originated from GM23338 treated with 0.5 ug/mL doxycycline hyclate for 4 days |
| 2951 | ENCFF996IOS | ENCSR486ZKB | CHIP:H3K36me3:neuron originated from H9 |
| 3282 | ENCFF869HFT | ENCSR592ETL | CHIP:H3K9ac:bipolar neuron originated from GM23338 treated with 0.5 ug/mL doxycycline hyclate for 4 days |
| 3358 | ENCFF205ZOQ | ENCSR619IUE | CHIP:CTCF:bipolar neuron originated from GM23338 treated with 0.5 ug/mL doxycycline hyclate for 4 days |
| 3559 | ENCFF424JDL | ENCSR683ORH | CHIP:H3F3A:bipolar neuron originated from GM23338 treated with 0.5 ug/mL doxycycline hyclate for 4 days |
| 3566 | ENCFF770SEF | ENCSR688OUZ | CHIP:H3K27me3:neuron originated from H9 |
| 3971 | ENCFF950LSJ | ENCSR814TSD | CHIP:H3K4me1:neuron originated from H9 |
| 3988 | ENCFF965UTU | ENCSR821GUG | CHIP:H4K20me1:bipolar neuron originated from GM23338 treated with 0.5 ug/mL doxycycline hyclate for 4 days |
| 3989 | ENCFF357TFM | ENCSR821IAK | CHIP:H3K4me2:bipolar neuron originated from GM23338 treated with 0.5 ug/mL doxycycline hyclate for 4 days |
| 4041 | ENCFF573DQZ | ENCSR839SFJ | CHIP:SMARCA4:bipolar neuron originated from GM23338 treated with 0.5 ug/mL doxycycline hyclate for 4 days |
| 4071 | ENCFF834UFV | ENCSR849YFO | CHIP:H3K4me3:bipolar neuron originated from GM23338 treated with 0.5 ug/mL doxycycline hyclate for 4 days |
| 4175 | ENCFF229IOE | ENCSR886KKK | CHIP:EZH2phosphoT487:bipolar neuron originated from GM23338 treated with 0.5 ug/mL doxycycline hyclate for 4 days |
| 4188 | ENCFF912TKR | ENCSR889GGV | CHIP:POLR2AphosphoS5:bipolar neuron originated from GM23338 treated with 0.5 ug/mL doxycycline hyclate for 4 days |
| 4245 | ENCFF346CWF | ENCSR905TYC | CHIP:H3K27ac:bipolar neuron originated from GM23338 treated with 0.5 ug/mL doxycycline hyclate for 4 days |
| 4316 | ENCFF588UZN | ENCSR928UVY | CHIP:H2AFZ:neuron originated from H9 |
| 4464 | ENCFF897UZP | ENCSR983CSB | CHIP:H2AFZ:bipolar neuron originated from GM23338 treated with 0.5 ug/mL doxycycline hyclate for 4 days |
| 80 | ENCFF025HHG | ENCSR000ENE | DNASE:brain microvascular endothelial cell |
| 81 | ENCFF353QSZ | ENCSR000ENF | DNASE:brain pericyte |
| 82 | ENCFF443IYY | ENCSR000ENG | DNASE:smooth muscle cell of the brain vasculature female |
| 179 | ENCFF735MKP | ENCSR026EOM | DNASE:brain female embryo (85 days) |
| 216 | ENCFF657KLL | ENCSR118WIQ | DNASE:brain embryo (112 days) |
| 240 | ENCFF064KBT | ENCSR156CLC | DNASE:brain female embryo (96 days) |
| 261 | ENCFF378OWK | ENCSR187PYY | DNASE:brain female embryo (142 days) |
| 319 | ENCFF536FNG | ENCSR309FOO | DNASE:brain female embryo (117 days) |
| 338 | ENCFF003FBE | ENCSR344FLH | DNASE:brain female embryo (109 days) |
| 370 | ENCFF947HEA | ENCSR420RWU | DNASE:brain male embryo (105 days) |
| 403 | ENCFF468SMI | ENCSR475VQD | DNASE:brain male embryo (72 days) and male embryo (76 days) |
| 421 | ENCFF482UKP | ENCSR507GFJ | DNASE:brain embryo (80 days) |
| 458 | ENCFF503ZTL | ENCSR572LDG | DNASE:brain male embryo (101 day) |
| 469 | ENCFF641HVS | ENCSR595CSH | DNASE:brain embryo (56 days) and male embryo (58 days) |
| 499 | ENCFF361UTR | ENCSR649KBB | DNASE:brain male embryo (122 days) |
| 524 | ENCFF225GCE | ENCSR706IDL | DNASE:midbrain male adult (78 years) and male adult (84 years) |
| 580 | ENCFF643ARA | ENCSR820XRX | DNASE:brain female embryo (105 days) |
| 644 | ENCFF622PZI | ENCSR947POC | DNASE:brain male embryo (104 days) |
| 1163 | ENCFF210HSY | ENCSR000DTA | CHIP:CTCF:brain microvascular endothelial cell |
| 1164 | ENCFF256KXI | ENCSR000DTC | CHIP:H3K4me3:brain microvascular endothelial cell |
| 1506 | ENCFF707FCC | ENCSR018OGF | CHIP:H3K9me3:brain female embryo (120 days) |
| 1739 | ENCFF614MKK | ENCSR094NYB | CHIP:H3K4me1:brain female embryo (120 days) |
| 1897 | ENCFF942CMQ | ENCSR143MZL | CHIP:H3K4me3:brain male embryo (122 days) |
| 1909 | ENCFF985OQZ | ENCSR146KXZ | CHIP:H3K9me3:brain female embryo (17 weeks) |
| 2623 | ENCFF321KUJ | ENCSR386JEL | CHIP:H3K4me1:brain female embryo (17 weeks) |
| 2916 | ENCFF139FPN | ENCSR479BPF | CHIP:H3K27me3:brain female embryo (120 days) |
| 3380 | ENCFF628RWE | ENCSR627STE | CHIP:H3K27me3:brain male embryo (122 days) |
| 3864 | ENCFF997IXW | ENCSR780FXX | CHIP:H3K4me3:brain female embryo (17 weeks) |
| 4015 | ENCFF495TLF | ENCSR829NJE | CHIP:H3K9me3:brain male embryo (122 days) |
| 4133 | ENCFF135VPT | ENCSR869WSC | CHIP:H3K36me3:brain female embryo (17 weeks) |
| 4182 | ENCFF988QYQ | ENCSR888JWS | CHIP:H3K36me3:brain male embryo (122 days) |
| 4509 | ENCFF204VPY | ENCSR997YTW | CHIP:H3K27me3:brain female embryo (17 weeks) |
| 4680 | CNhs10617 | NA | CAGE:brain, adult, pool1 |
| 4980 | CNhs11796 | NA | CAGE:brain, adult, |
| 4981 | CNhs11797 | NA | CAGE:brain, fetal, pool1 |
| 362 | ENCFF083ELT | ENCSR401ESD | DNASE:tibial nerve female adult (51 year) |
| 543 | ENCFF094FZF | ENCSR754WNA | DNASE:tibial nerve male adult (37 years) |
| 1931 | ENCFF314DQL | ENCSR154GUK | CHIP:POLR2A:tibial nerve female adult (51 year) |
| 1944 | ENCFF134TOV | ENCSR157ZVP | CHIP:H3K36me3:tibial nerve female adult (53 years) |
| 2408 | ENCFF921FBI | ENCSR314SPW | CHIP:H3K4me3:tibial nerve female adult (53 years) |
| 2782 | ENCFF725SHA | ENCSR434XLP | CHIP:CTCF:tibial nerve male adult (37 years) |
| 2892 | ENCFF423FWZ | ENCSR469POZ | CHIP:CTCF:tibial nerve female adult (53 years) |
| 2918 | ENCFF851LAU | ENCSR479GPU | CHIP:H3K9me3:tibial nerve female adult (53 years) |
| 3212 | ENCFF004UZT | ENCSR569SZK | CHIP:EP300:tibial nerve female adult (51 year) |
| 3246 | ENCFF469CEQ | ENCSR580XUK | CHIP:EP300:tibial nerve male adult (37 years) |
| 3338 | ENCFF338MNH | ENCSR611YUJ | CHIP:H3K27me3:tibial nerve female adult (53 years) |
| 3836 | ENCFF420PYG | ENCSR771YJT | CHIP:H3K27ac:tibial nerve female adult (53 years) |
| 3994 | ENCFF991GMN | ENCSR822PJT | CHIP:CTCF:tibial nerve female adult (51 year) |
| 4075 | ENCFF486TEG | ENCSR850RVA | CHIP:H3K4me1:tibial nerve female adult (53 years) |
| 4330 | ENCFF188HNM | ENCSR935XOT | CHIP:POLR2A:tibial nerve female adult (53 years) |
| 5230 | CNhs13449 | NA | CAGE:optic nerve, |

**Supplementary Table 4**. The human brain cerebral cortex is divided into two cerebral hemispheres. Each hemisphere is divided into four lobes: frontal, parietal, temporal and occipital. We focus on the 29 brain regions of interest (ROIs) for each hemisphere in the above four brain lobes.

| Brain lobe | ROI | Full name |
| --- | --- | --- |
| Temporal lobe | bankssts | Banks of the superior temporal sulcus |
| Temporal lobe | entorhinal | Entorhinal cortex |
| Temporal lobe | fusiform | Fusiform gyrus |
| Temporal lobe | inferiortemporal | Inferior temporal gyrus |
| Temporal lobe | middletemporal | Middle temporal gyrus |
| Temporal lobe | parahippocampal | Parahippocampal cortex |
| Temporal lobe | superiortemporal | Superior temporal gyrus |
| Temporal lobe | temporalpole | Temporal pole |
| Temporal lobe | transversetemporal | Transverse temporal gyrus |
| Frontal lobe | caudalmiddlefrontal | Caudal middle frontal |
| Frontal lobe | lateralorbitofrontal | Lateral orbitofrontal cortex |
| Frontal lobe | medialorbitofrontal | Medial orbitofrontal cortex |
| Frontal lobe | paracentral | Paracentral lobule |
| Frontal lobe | parsopercularis | Pars opercularis |
| Frontal lobe | parsorbitalis | Pars orbitalis |
| Frontal lobe | parstriangularis | Pars triangularis |
| Frontal lobe | precentral | Precentral gyrus |
| Frontal lobe | rostralmiddlefrontal | Rostral middle frontal gyrus |
| Frontal lobe | superiorfrontal | Superior frontal gyrus |
| Frontal lobe | frontalpole | Frontal pole |
| Occipital lobe | cuneus | Cuneus |
| Occipital lobe | lateraloccipital | Lateral occipital sulcus |
| Occipital lobe | lingual | Lingual gyrus |
| Occipital lobe | pericalcarine | Pericalcarine cortex |
| Parietal lobe | inferiorparietal | Inferior parietal lobe |
| Parietal lobe | postcentral | Postcentral gyrus |
| Parietal lobe | precuneus | Precuneus |
| Parietal lobe | superiorparietal | Superior parietal lobule |
| Parietal lobe | supramarginal | Supramarginal gyrus |
